# Polygenic and environmental determinants of tics in the Avon Longitudinal Study of Parents and Children cohort

**DOI:** 10.1101/2021.05.10.21256958

**Authors:** Mohamed Abdulkadir, Jay A. Tischfield, Gary A. Heiman, Pieter J. Hoekstra, Andrea Dietrich

## Abstract

**BACKGROUND:** Tourette syndrome (TS) is caused by multiple genetic and environmental factors. Yet, little is known about the interplay of these factors in the occurrence of tics in the general population.

**METHODS:** Using logistic regression, we investigated whether polygenic risk score (PRS) of TS and pregnancy-related environmental factors together enhance the explained variance of tic occurrence (as opposed to separate analysis) in the Avon Longitudinal Study of Parents and Children. We included a cumulative adverse pregnancy risk score, maternal anxiety and depression, and maternal smoking and alcohol use during pregnancy. We investigated possible independent (i.e. additive) genetic and environmental effects, gene-environment correlations (*r*GE), gene-environment interactions (G x E), and mediation effects in explaining tic presence.

**RESULTS:** Models that contained the PRS and the cumulative adverse pregnancy risk score, maternal anxiety, or maternal depression (but not maternal smoking and alcohol use) explained significantly more variance of tic presence compared to models including only the PRS, pointing to additive effects. Furthermore, we found that maternal anxiety, depression, and smoking were mediated by the cumulative adverse pregnancy risk score, and were thus all indirectly associated with tics through pregnancy complications. We did not find *r*GE or G x E.

**CONCLUSIONS:** We found evidence for both direct and indirect associations of environmental risk factors in relation to tics in the general population. Combining PRS and environmental risk factors improve our understanding of tics compared to considering these factors in isolation, suggesting both additive and mediation effects.

## INTRODUCTION

Tourette syndrome (TS) is a polygenic disorder with an estimated population-based heritability of approximately 77% (1). A substantial portion of this heritability can be captured through the use of common variants; the single nucleotide polymorphism (SNP)-based heritability of TS is estimated at 58% (2). Yu et al. and our group independently demonstrated that an aggregate score of common variants identified in a TS GWAS, referred to as polygenic risk score (PRS), is significantly associated with a range of tic traits in the general population (3,4). These findings (2–4), together with evidence from epidemiological studies (5–7), encourage the conceptualization of tic disorders as part of a phenotypic continuum with TS on the extreme end of the spectrum and milder or less chronic tics (e.g., transient tics) on the other end. This notion could imply that TS could be studied more effectively by also including milder tic phenotypes that are more prevalent; the lifetime prevalence of TS is between 0.32 and 0.85% while the prevalence of transient tics is estimated between 1.60 and 5.61% (8,9).

While twin and family studies suggest substantial heritability of tic disorders, there is still an unexplained phenotypical variance of tic disorders that could partly be attributed to environmental factors (10–12). Environmental factors may have an independent contribution, but also be a resultant of an underlying genetic risk (13). Recent larger prospective studies provide clear evidence for the involvement of pregnancy related risk factors in TS (14–17), albeit with variable results. The discrepancy in results between studies (14,15) may perhaps be partly explained by unaccounted genetic factors, rarely explored together with environmental factors in TS research so far. One exception is a large Swedish population-based birth cohort study (N = 3,026,861) using a full sibling design, in which the authors reported that a cumulative score of perinatal risk factors was related to TS, largely independent from unmeasured environmental and genetic confounding, suggesting causal influences (14). At the same time, the study found no evidence for prenatal maternal smoking after controlling for this confounding (14). This is in contrast with the findings from another large prospective study, a Danish National Birth Cohort (N = 73,073), which implicated prenatal maternal smoking in TS, yet did not account for a genetic contribution (15).

Maternal mental health during and after pregnancy also appear to be important environmental factors and have been implicated in other psychiatric disorders (18) as well as in tic disorders. That is, in a prospective general population cohort, the Avon Longitudinal Study of Parents and Children cohort (ALSPAC; (19,20)) a role for maternal anxiety and depression during pregnancy and the first 8 months after giving birth has been implicated in tic disorders (16). Taken together, findings from these prospective studies (14–16) suggest involvement of pregnancy related risk factors in tic disorders, but also emphasize the need of including a genetic component; the association between pregnancy related risk factors and tic disorders could be due to shared genetic liability. Furthermore, risk factors during pregnancy (15,16) may be related with each other implying possible mediation effects; e.g. mothers who experience anxiety or depression during pregnancy may be more likely to smoke during pregnancy (21–25), which in turn may be associated with tics. In addition, maternal smoking has been related to pregnancy adversities, such as gestational diabetes (26), which again may imply a mediation effect; mothers who smoke during pregnancy may experience more pregnancy complications, which in turn may be associated with tic presence in their offspring.

To date, the precise contribution and interplay of genetic and environmental factors in tic disorders has remained largely unexamined (10). PRS have been applied in gene-environment interaction (G x E) studies of major depressive disorder and schizophrenia (27–29) but not in TS. In the current study, we aimed to investigate how previously identified pregnancy related risk factors (i.e., a cumulative score of adverse pregnancy risk factors, and maternal anxiety, depression, smoking and alcohol use) together with a PRS derived from a TS GWAS are associated with the presence of tics in ALSPAC (19,20). We were specifically interested whether genetic and environmental effects were independent (i.e., showed additive effects) or correlated, and/or showed interaction or mediation effects in their association with tics. Based on the broader literature of neuropsychiatric disorders, we expected to find associations at various levels.

## METHODS AND MATERIALS

### Participants

The current study included 612 adolescents with tics and 4,201 controls from ALSPAC (19,20,30), both with genotype and phenotype data available at age 13.8 years. The ALSPAC study is an ongoing population-based birth cohort study of mothers and their children (that were born between 1 April 1991 and 31 December 1992) residing in the southwest of England (UK). From the 14,541 pregnancies, 13,988 were alive at 1 year. At age 7 years, this sample was bolstered with an additional 913 children. The total sample size for analyses using any data collected after the age of seven is therefore 15,454 pregnancies; of these 14,901 were alive at 1 year of age. Participants are assessed in regular intervals from birth, using clinical interviews, self-reported questionnaires, medical records, and physical examinations. We did not exclude participants based on IQ nor the presence of autism spectrum disorder. The study website contains details of available data through a fully searchable data dictionary: http://www.bristol.ac.uk/alspac/researchers/our-data/.

### Ethical approval

Ethical approval was obtained from the ALSPAC Ethics and Law Committee. All participants provided written informed consent or assent. Consent for biological samples has been collected in accordance with the Human Tissue Act (2004). Informed consent for the use of data collected via questionnaires and clinics was obtained from participants following the recommendations of the ALSPAC Ethics and Law Committee at the time.

### Phenotypic assessment

The primary outcome measure was the presence of tics, either motor and/or vocal, occurring at least once a week at age 13.8 years as reported by the mother; individuals fulfilling these criteria were coded as 1 and those not fulfilling these criteria as 0; mothers reported whether their child exhibited in the past year repeated movements of the 1) face and head, 2) neck, shoulder, or trunk, 3) arms, hands, legs, feet; and had 4) repeated noises and sounds or 5) repeated words or phrases; (“not at all” =0, “definitely” and “probably” = 1). The frequency of these tic symptoms was also assessed (“less than once a month” =1; “1–3 times a month” =2; “about once a week” =3; “more than once a week” =4; “every day” = 5). Our outcome measure for tic presence is consistent with the ‘Tourette syndrome/chronic tic disorder broad’ definition of Scharf et al. (31). We chose to only include this definition of tics over our previously reported tic phenotypes (“Tics intermediate”, “Tics all”) (4,31) as the explained variance by the TS PRS was the highest for this tic phenotype (4).

### Environmental predictors

#### Cumulative adverse pregnancy risk score

We included 15 pregnancy related variables summarized into 9 distinct exposures, given that several variables informed about the same exposure but at different time-points (e.g., maternal infection; see Table S3). The absence of exposure to the pregnancy related variable was coded as 0 and presence of exposure was coded as 1. Then, a cumulative adverse pregnancy risk score was constructed from all 9 exposures (possible range: 0-9; Table S3). Note that the maternal anxiety and depression variables, and the maternal substance use variables (as discussed below) were not part of the cumulative score of pregnancy factors as we intended to investigate the effects of these variables separately.

#### Maternal anxiety

Mothers completed the self-report anxiety subscale of the Crown-Crisp Experiential Index (8 items (30,32,33)) at three time-points (prenatally at 18 weeks and postnatally at 8 weeks and 8 months) (Table S1). For each time-point, the maternal total anxiety score was dichotomized (0 = score in the bottom two tertiles, 1 = score in the top tertile) (16). Presence of maternal anxiety was defined as being scored in the top tertile at all three time-points, consistent with a previous ALSPAC paper (16). For the mediation analyses we only utilized the Crown-Crisp Experiential Index score measured 18 weeks prenatal referred to as prenatal maternal anxiety, since we explored associations with other prenatal factors.

#### Maternal depression

Analogous to the anxiety subscale of the Crown-Crisp Experiential Index (30,32,33), mothers reported on the 8-item depression subscale at three time-points (prenatally at 18 weeks and postnatally at 8 weeks and 8 months) (Table S2). Similarly, presence of maternal depression was defined as being scored in the top tertile on the total depression score at all three time-points and for the mediation analyses we limited to the analyses to the Crown-Crisp Experiential Index measured at 18 weeks prenatal (16).

#### Maternal substance use

In accordance with a previous ALSPAC publication (17), we examined the presence of exposure to respectively maternal smoking and alcohol drinking during the last two months of the pregnancy; we did not include maternal cannabis use in our analyses as this exposure was infrequently endorsed (N < 100). For each variable, presence of exposure was coded as 1 and the absence thereof as 0.

### Confounders

Based on previous studies, the child’s sex, primiparity (being first-born), maternal age (at time of the birth of their child), and socioeconomic status (SES) were included as confounders (12,17,34). Maternal SES was based on the occupation of the mother and was measured during and after pregnancy up to 3 years postnatally (34). We also calculated the first four ancestry-informative principal components to account for potential residual population stratification.

### PRS

#### Genotyping

Genotyping data was available of 9,915 children out of the total of 14,541 ALSPAC participants. Genotyping and the necessary quality control steps undertaken to clean the data is described in more detail in a previous study (4). After removal of SNPs with excessive missingness (i.e., call rate less than 95%), minor allele frequency less than 1%, a departure from the Hardy–Weinberg equilibrium (P value < 5 x 10-7), and an Impute2 information quality of metric of < 0.8; a total of 8,941 individuals (4,580 males, 4,361 females) and 6,976,085 SNPs remained eligible for analyses.

#### PRS

Details on how the PRS were derived were previously described (4). Briefly, PRS were calculated using PRSice V2.0.7.beta (35) and were based on the alleles and effect sizes reported in the second GWAS of TS (3). A PRS was calculated for each individual as a sum of the risk alleles they carried weighted by the odds ratio reported in the second GWAS of TS (3). The PRS were based on all available SNPs and we report the PRS model with the most predictive *P*-value thresholds as measured by Nagelkerke’s R^2^.

### Statistical analyses

Statistical analyses were carried out using R (version 3.5.0) statistical software tool (36).

#### Univariate analyses

We first conducted univariate analyses to individually assess whether each single predictor (PRS and all four environmental exposures) as well as each potential confounder (i.e., child’s sex, maternal age, primiparity, and SES) was associated with the presence of tics using logistic regression analyses comparing cases versus controls. Variables that were not related to tic presence (*P* ≥ 0.05) were excluded from further analyses. We also carried out gene-environment correlations (*r*GE) in which we tested whether the PRS was related to the environmental variables using logistic regression analyses with a significance threshold set at *P* < 0.05. Univariate analyses were not corrected for multiple testing as they served only for the selection of variables for the multivariate analyses.

#### Multivariate analyses

First, to test for the additive effects of genetic and environmental influences, we investigated logistic regression models for each of the four environmental exposures (cumulative adverse pregnancy risk score, maternal anxiety, maternal depression, maternal smoking and alcohol use) separately in case of a significant (*P* < 0.05) univariate result, entering the PRS, potential confounders, the first four ancestry-informative principal components, and the respective environmental variable into one model and compared this with a model that only contained the PRS and the relevant confounders (referred to as the reference model), using the likelihood ratio test in R (36). We also explored a full model, where we entered the PRS, potential confounders, all significant environmental variables and compared that model to the reference model. We report both R^2^ and the area under the curve (AUC) to evaluate variance explained (by the model) and model fitness, respectively. We also report the significance of each predictor within the full model using a logistic regression model.

Second, we tested for each environmental exposure, whether there was a G x E interaction effect between the PRS and the environmental variable adding the interaction term to the model. In case of a significant *r*GE (see above), the residuals were included to this interaction model (37). Correction for multiple comparisons between the models was done using the Benjamini-Hochberg false discovery rate (FDR) method. The significance threshold was met if the FDR adjusted empirical *P* value (i.e. *Q*) was < 0.05.

#### Mediation analyses

Since the literature supports an association between maternal anxiety/depression symptoms and maternal smoking (i.e., anxiety/depression is associated with higher odds of smoking) (23–25), we tested whether maternal smoking mediates the effect of these two variables on tic presence. Previous research also suggests that mothers who experience anxiety or depression during pregnancy have an increased risk for perinatal complications (38–41). Therefore, we also sought to understand whether the cumulative adverse pregnancy risk score mediates between maternal anxiety/depression and tic presence. For our mediation analyses of maternal anxiety/depression we only included the prenatal (at 18 weeks) Crown-Crisp Experiential Index scores. Furthermore, we investigated whether the effects of maternal smoking were mediated through the cumulative adverse pregnancy risk score, since studies reported that maternal smoking increases the odds of pregnancy complications (42–45). Finally, in case of a significant association between the PRS and the environmental variables, we planned mediation models to test whether the association of PRS with tics are mediated by environmental factors. These mediation analyses were considered as exploratory to better understand possible interrelationships between the environmental variables; hence we did not apply correction for multiple testing and set the threshold of significance at *P* < 0.05. All mediation analyses were carried out with the mediation package in R (46). Direct (average direct effect, ADE), indirect effects (average causal mediation effect, ACME), and total effect (sum of ADE and ACME) were estimated controlling for the child’s sex, maternal age, primiparity, SES, and in case of a significant *r*GE also the TS PRS(47). As measure of the effect size of the mediation effect we report the proportion mediated which is calculated by dividing the ACME by the total effect.

## RESULTS

### Sample description

Table 1 presents the clinical characteristics of the ALSPAC participants of whom both genotype and phenotype data were available at age 13.8 years.

**Table 1:** Clinical characteristics and univariate analysis of the environmental risk factors of participants in the Avon Longitudinal Study of Parents and Children study

| | Cases (N = 612)<sup>a</sup> | | Controls (N = 4,201)<sup>a</sup> | | $\beta$ | Z | OR (95% CI) | *P*<sup>b</sup> |
| --- | --- | --- | --- | --- | --- | --- | --- | --- |
|  | N | % | N | % |  |  |  |  |
| Sex (% male) | 395 | 64.5 | 2004 | 47.7 | 0.69 | 7.68 | 1.99(1.67-2.38) | **1.95 x 10<sup>-14</sup>** |
| Primiparity (firstborn) | 325 | 53.1 | 1855 | 44.2 | 0.25 | 4.07 | 1.29(1.13-1.49) | **4.62 x 10<sup>-5</sup>** |
| Maternal anxiety<sup>c</sup> | 88 | 14.4 | 332 | 7.9 | 0.67 | 5.24 | 1.96(1.52-2.52) | **1.58 x 10<sup>-7</sup>** |
| Maternal depression<sup>c</sup> | 84 | 13.7 | 385 | 9.16 | 0.45 | 3.52 | 1.57(1.22-2.01) | **4.37 x 10<sup>-4</sup>** |
| Maternal smoking<sup>d</sup> | 102 | 16.7 | 512 | 12.2 | 0.38 | 3.20 | 1.46 (1.15 – 1.84) | **0.001** |
| Maternal alcohol<sup>d</sup> | 310 | 50.6 | 2251 | 53.6 | -0.09 | -1.10 | 0.91(0.76-1.08) | 0.27 |
| | Mean (SD) | Observed range | Mean (SD) | Observed range | $\beta$ | Z | OR (95% CI) | *P*<sup>b</sup> |
| Maternal SES<sup>e</sup> | 6.23 (2.09) | 1.2-15 | 6.0(1.99) | 1.2-15 | 0.06 | 2.58 | 1.06(1.01-1.11) | **0.01** |
| Maternal age | 29.05(4.57) | 17-42 | 29.38(4.45) | 15-45 | -0.02 | -1.66 | 0.98(0.96-1.0) | 0.09 |
| Cumulative adverse pregnancy risk score<sup>f</sup> | 2.09(1.04) | 0-5 | 1.98(0.99) | 0-6 | 0.10 | 2.19 | 1.11 (1.01 – 1.22) | **0.028** |

SES, socioeconomic status. PRS, polygenic risk score
<sup>a</sup>Cases were defined as individuals who have experienced parent-reported motor and vocal tics at least once a week at age 13.8 years. Individuals not fulfilling these criteria were considered controls.
<sup>b</sup>Univariate analyses were carried out using logistic regressions comparing cases and controls. The significance threshold was set at $P < 0.05$ . Only variables with $P < 0.05$ (in bold for clarity purposes) were included in the multivariate analyses (Table 2).
<sup>c</sup>Maternal anxiety and maternal depression were measured using the self-rated Crown-Crisp Experiential Index (30,32,33).
<sup>d</sup>Self-report (yes/no) on maternal smoking and alcohol drinking during the last two months of the pregnancy.
<sup>e</sup>Maternal socioeconomic status derived from occupation data assessed at 18 weeks' gestation. A higher score indicates a lower SES.
<sup>f</sup>A cumulative adverse pregnancy risk score constructed from 15 prenatal risk factors (Table S3).

### Univariate analyses

#### Confounders and gene-environment correlations

Male sex of the child, lower maternal SES, and being firstborn (primiparity) were significantly associated with presence of tics (Table 1). Hence, these variables were included in the multivariate regressions as confounders. Maternal age was unrelated to tics and therefore not included. There were no significant *r*GE between the PRS and any of the environmental variables as determined by logistic regression (Table S4).

#### Univariate associations between the environmental factors and tic presence

*Polygenic risk score of TS*. The significant association between the PRS based on the TS GWAS and the presence of tics is reported in our previous work (4).

#### Cumulative adverse pregnancy risk score

A higher cumulative score of adverse pregnancy risk factors was significantly associated with tic presence in the univariate analysis (*P* = 0. 028; OR = 1.11; 95% CI = 1.01 – 1.22; Table 1).

#### Maternal anxiety and depression

Separate analysis of maternal anxiety (*P* = 1.58 × 10^−7^; OR = 1.96; 95% CI = 1.52 – 2.52) and maternal depression (*P* = 4.37 × 10^−4^; OR = 1.57; 95% CI = 1.22 – 2.01) showed that both were significantly associated with tic presence; mothers with anxiety and depression scores in the top tertile during pregnancy were more likely to have an offspring with tics (Table 1).

#### Maternal substance use

Maternal smoking during pregnancy was significantly associated with tic presence in the univariate analysis (*P* = 0.001; OR = 1.46; 95% CI = 1.15 – 1.84). We found no evidence of a univariate association of maternal alcohol with tic presence, therefore this variable was not included in subsequent multivariate analyses (Table 1).

### Multivariate analyses

#### Cumulative adverse pregnancy risk score

The model containing the PRS and the cumulative adverse pregnancy risk score explained significantly more variance (PRS + cumulative adverse pregnancy risk score model R^2^ = 0.050; AUC = 0.638; Q = 0.004; Table 2) of tic presence as compared to the reference model. This suggests additive effects of the environmental variable above an independent genetic contribution. We found no evidence for a significant G x E interaction between the PRS and cumulative adverse pregnancy risk score.

**Table 2:** Multivariate logistic regression models of tic presence in the Avon Longitudinal Study of Parents and Children study

| Models (N = 2,192)<sup>a</sup> | Terms in model | R<sup>2</sup> | AUC | Likelihood ratio test ($\chi^2$) | P (comparison to reference model)<sup>b</sup> | Q<sup>c</sup> |
| --- | --- | --- | --- | --- | --- | --- |
| Reference model | PRS | 0.043 | 0.628 |  |  |  |
| Maternal anxiety<sup>d</sup> | PRS, maternal anxiety | 0.055 | 0.644 | 15.12 | 0.0001 | **0.0003** |
| Maternal depression<sup>d</sup> | PRS, maternal depression | 0.049 | 0.635 | 7.24 | 0.007 | **0.009** |
| Cumulative adverse pregnancy risk score<sup>e</sup> | PRS, cumulative adverse pregnancy risk score | 0.050 | 0.638 | 9.10 | 0.003 | **0.004** |
| Maternal smoking<sup>f</sup> | PRS, maternal smoking | 0.046 | 0.635 | 3.78 | 0.052 | 0.052 |
| Full model | PRS, maternal anxiety, maternal depression, prenatal risk score, maternal smoking | 0.063 | 0.654 | 24.31 | 6.9 x 10<sup>-5</sup> | **0.0003** |

PRS, polygenic risk score
<sup>a</sup>Sample size reflects individuals with complete data on the environmental variables, genetic data, and confounder variables.
<sup>b</sup>All models included sex, maternal socioeconomic status, and primiparity that were related to tic presence in our univariate analyses (Table 1). In addition, the models were adjusted for the first four ancestry-informative principal components.
<sup>c</sup>Correction for multiple comparisons between the models was done using the Benjamini-Hochberg false discovery rate (FDR) method. The significance threshold was met if the FDR adjusted *P* value (i.e. *Q*) was < 0.05 (in bold for clarity purposes).
<sup>d</sup>Maternal anxiety and maternal depression were measured using the self-rated Crown-Crisp Experiential Index (30,32,33).
<sup>e</sup>A cumulative score constructed from 15 prenatal risk factors (Table S3).
<sup>f</sup>Self-report (yes/no) on maternal smoking during the last two months of the pregnancy.

#### Maternal anxiety and depression

For maternal anxiety, we observed that the model containing both the PRS and maternal anxiety (PRS + maternal anxiety model R^2^= 0.055; AUC = 0.644; Q = 0.0003) significantly explained more variance of tic presence compared to the reference model (R^2^ = 0.043, AUC = 0.628) that contained only the PRS (Table 2). The model containing maternal depression and the PRS also significantly explained more variance (PRS + maternal depression model R^2^= 0.049; AUC = 0.635; Q = 0.009) of tic presence as compared to the reference model. We found no significant G x E interaction effects between the PRS and respectively maternal anxiety and depression.

#### Maternal smoking

The multivariate model that included the PRS and maternal smoking did not significantly explain more variance of tic presence as compared to the reference model (R^2^ PRS + maternal smoking = 0.046 and AUC = 0.635; Q = 0.052; Table 2). There was no significant G x E interaction between the PRS and maternal smoking.

#### Full model

Finally, all environmental variables were entered into one full model together with the PRS and the relevant confounders. This model significantly explained more variance of tic presence compared to a model containing only the PRS (full best-fitting model R^2^= 0.063; AUC = 0.654; Q = 0.0003; Table 2). Within this multivariate model the PRS, maternal anxiety, and the cumulative adverse pregnancy risk score showed an independent significant association with tic presence, but not maternal depression and maternal smoking (Table 3).

**Table 3:**
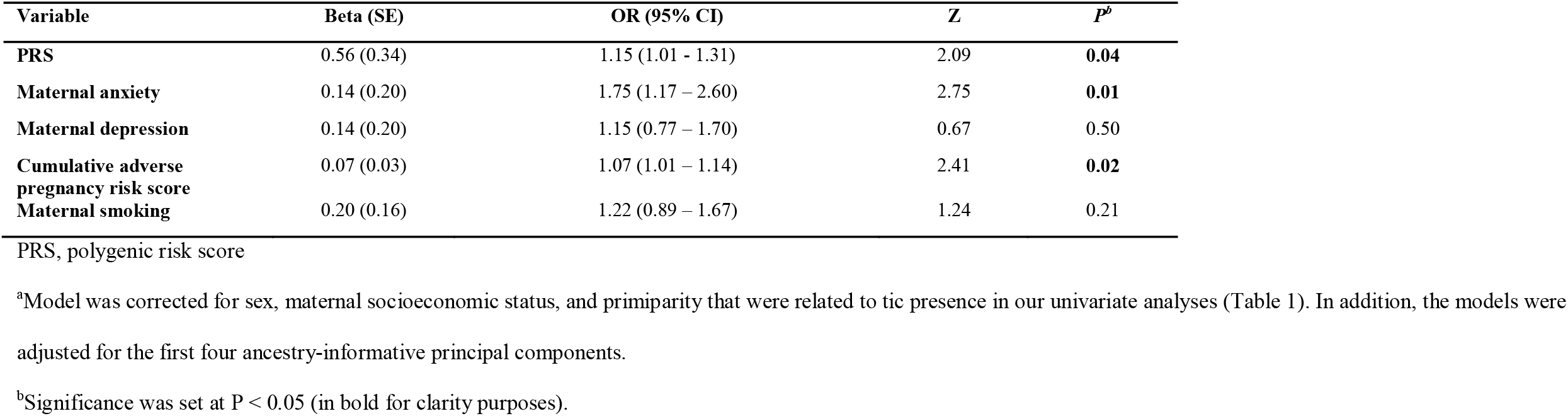
Full model containing all environmental variables and the polygenic risk score explaining tic presence in the Avon Longitudinal Study of Parents and Children study^a^

### Exploratory mediation analyses

Given the absence of *r*GE, TS PRS was not included in mediation models as a variable of interest or as a potential confounder.

#### Relation between prenatal maternal anxiety/depression and tic presence as mediated by the cumulative adverse pregnancy risk score

We observed that the cumulative adverse pregnancy risk score significantly mediated the association between prenatal maternal anxiety and tic presence (ACME = 0.0005; *P* = 0.01; Figure 1 & Table 4); i.e., mothers who reported anxiety during the pregnancy of their child were more likely to experience more complications during the pregnancy, which in turn were associated with tics in their offspring. A similar significant effect in the same direction was found for prenatal maternal depression (ACME = 0.003; *P* = 0.004).

**Table 4:** Exploratory causal mediation analyses predicting tic presence in the Avon Longitudinal Study of Parents and Children study^a^

| Predictor | Mediator | Proportion mediated | ACME (95% CI) | *P* ACME<sup>b</sup> | ADE (95% CI) | *P* ADE<sup>b</sup> |
| --- | --- | --- | --- | --- | --- | --- |
| Prenatal maternal anxiety | Cumulative adverse pregnancy risk score | 0.08 | 0.0005 (0.0001 - 0.001) | **0.01** | 0.006 (0.003 - 0.01) | **<0.001** |
| Prenatal maternal depression | Cumulative adverse pregnancy risk score | 0.12 | 0.003 (0.0009 – 0.01) | **0.004** | 0.02 (0.01 - 0.04) | **0.002** |
| Prenatal maternal anxiety | Maternal smoking | 0.038 | 0.001 (-0.0003 - 0.0001) | 0.15 | 0.025 (0.015 – 0.04) | **<0.001** |
| Prenatal maternal depression | Maternal smoking | 0.06 | 0.001 (-0.0002 - 0.0001) | 0.10 | 0.02 (0.011 - 0.03) | **<0.001** |
| Maternal smoking | Cumulative adverse pregnancy risk score | 0.27 | 0.009 (0.003 - 0.02) | **0.006** | 0.03 (-0.01 - 0.06) | 0.18 |

ACME, average causal mediation effect; ADE, average direct effect.
<sup>a</sup>All models included sex, maternal socioeconomic status, and primiparity that were related to tic presence in our univariate analyses (Table 1).
<sup>b</sup>The significance threshold was met if *P* value was < 0.05 (bold).

**Figure 1:**
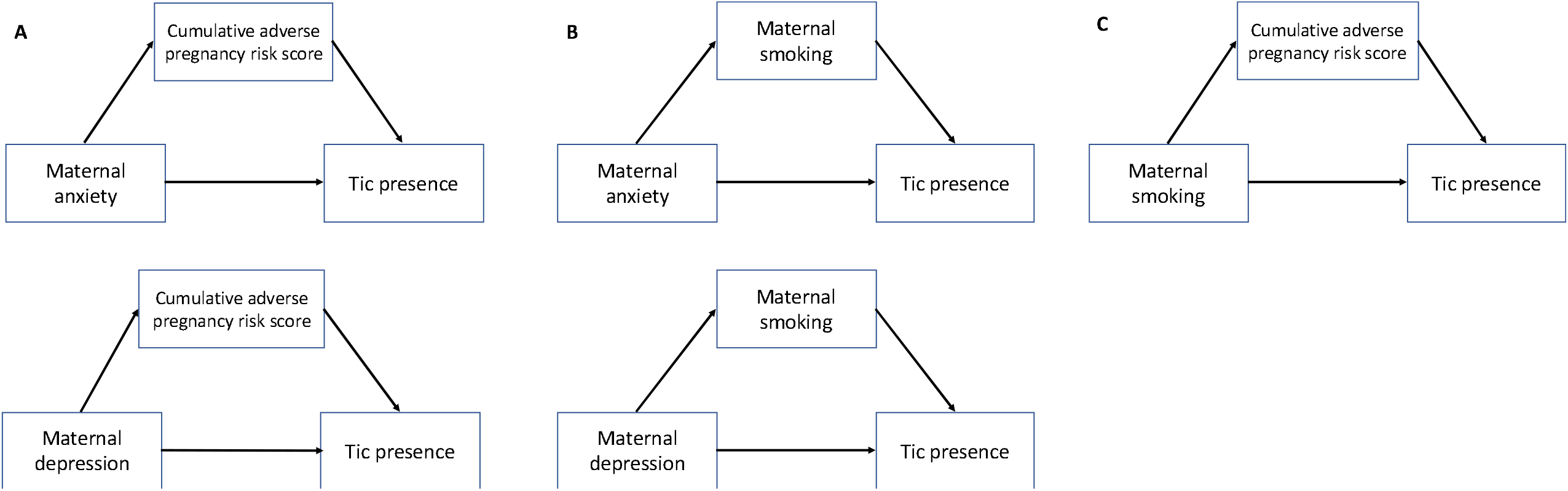
Directed Acyclic Graph (DAG) to depict an overview of the mediation analyses. **A**. Testing whether the association between prenatal maternal anxiety/depression and tic presence is mediated through the cumulative adverse pregnancy risk score; i.e., whether prenatal maternal anxiety/depression during pregnancy can lead to a higher cumulative adverse pregnancy risk score that in turn can increase the odds of tics in their offspring. **B**. Testing whether the association between prenatal maternal anxiety/depression is mediated through maternal smoking during pregnancy; i.e., whether mothers that experience maternal anxiety/depression are more likely to smoke during pregnancy which in turn can increase the odds of tics in their offspring. **C**. Testing whether the association between maternal smoking and tic presence is mediated by the cumulative adverse pregnancy risk score; i.e., whether maternal smoking during pregnancy can lead to more pregnancy complications (an increase in the cumulative adverse pregnancy risk score) that in turn can increase the odds of tics in their offspring.

#### Relation between prenatal maternal anxiety/depression and tic presence as mediated by maternal smoking

The association between prenatal maternal depression/anxiety and tic presence was not significantly mediated by maternal smoking (Figure 1 & Table 4).

#### Relation between maternal smoking and tic presence mediated by the cumulative adverse pregnancy risk score

The effect of maternal smoking on tic presence was significantly mediated by the cumulative adverse pregnancy risk score (ACME = 0.009, *P* = 0.006; Figure 1 & Table 4); i.e., mothers who smoked during pregnancy were at an increased risk of experiencing more pregnancy complications which in turn was associated with tic presence in their offspring.

## DISCUSSION

We studied the contribution of environmental risk factors during pregnancy (a cumulative adverse pregnancy risk score, and maternal anxiety, depression, smoking and alcohol use) and genetic factors (PRS based on a GWAS of cases with TS) to the presence of tics in adolescents from the large ALSPAC population cohort. Our study demonstrates that TS PRS and pregnancy related environmental risk factors together have greater explanatory power than either of these factors alone in relation to tic occurrence. However, we found no evidence of *r*GE or G x E between the PRS and the environmental risk factors. The findings from this study thus point towards additive effects; both genetic and environmental factors may exist alongside each other explaining tic occurrence.

The environmental risk factors investigated in this population study have previously been related to clinical cases of TS (for a review see (10)). Similarly, recent new evidence has indicated a shared genetic basis of clinically defined TS and a more broadly defined tic phenotype (as used in our study, being more prevalent in the general population) (3,4). Our study supports that next to shared genetic also shared environmental mechanisms are underlying tics, considered as part of one spectrum from non-clinical to clinical levels.

In line with previous findings in clinical samples (14), we found an association between the number of pregnancy complications and the presence of tics within the general population. While previous studies investigated a broader range of pre- and perinatal factors, in this study we focused on prenatal factors associated with mothers’ poorer medical and mental health (such as high blood pressure, infections or medication use and anxiety/depression, substance use). The current study expands on these previous findings and suggests that cumulative pregnancy complications are also associated with the broader spectrum of tic phenotypes as present in the general population.

Furthermore, we found direct effects of maternal anxiety and depression on tics, consistent with a previous ALSPAC study using a smaller sample of individuals with a phenotype approximating TS (16). Interestingly, the association between maternal anxiety and depression and tics appeared in part to be mediated by the cumulative adverse pregnancy risk score. That is, mothers who experience higher levels of anxiety or depression during pregnancy are more likely to experience (more) pregnancy complications, which in turn are associated with the occurrence of tics in their offspring.

After controlling for confounders (i.e., sex, SES, primiparity, other environmental risk factors and the TS PRS) maternal smoking during pregnancy was not directly associated with tics. This is in agreement with findings from a previous ALSPAC study (16) by Mathews et al. using a multivariable analysis, and a Swedish population-based cohort study by Brander et al. (14) that controlled for genetic confounding using a sibling design. Similar studies on attention deficit hyperactivity disorder (often comorbid to TS) using a genetically sensitive design also suggested no association with maternal smoking after controlling for unmeasured familial factors (i.e., shared genetics and/or family environment) (48,49). Whether maternal smoking contributes to TS risk still is debated in the literature (14,15,17,50,51). Nevertheless, our mediation analyses suggest that maternal smoking could indirectly be related to tics through cumulative pregnancy aversities. Note that this mediation effect was independent from TS PRS, as we did not observe *r*GE in our study.

An important finding to highlight from our analyses is that the number of pregnancy complications seems to play a central role; it is directly associated with tics but also, as discussed above, mediates the associations between the other investigated environmental risk factors (anxiety, depression, and smoking) and tics. The observed association between pregnancy complications with maternal anxiety/depression and maternal smoking is supported by previous findings (26,39,52).

A few strengths and limitations should be noted. While the use of the second TS GWAS for calculation of the PRS (3) is a strength, it should be noted that current PRS explain only a very small proportion of the phenotype. Greater predictive power will be expected in the future with larger GWAS and the discovery of rare gene variants also involved in tic disorders (53–55). Another asset is the use of the large ALSPAC sample (19,20,30,33), prospectively assessing prenatal risk factors collected at different time-points during pregnancy and allowing for analysis of a broader tic phenotype occurring in the general population, supporting generalizability of previous findings based on clinical cases. Despite the large sample, still specific prenatal complications may have been too infrequent to be studied in isolation, yet our study affirms the value of using a cumulative score of pregnancy adversities. Another challenge is the widely diverse selection of pre- and/or postnatal variables across studies; by focusing on pregnancy we aimed to study a more parsimonious group. Moreover, the ALSPAC sample is homogenous in terms of ancestry making it an ideal target population to study genetic risk factors. Yet, the current sample and TS PRS may still yield insufficient power to detect G x E or *r*GE with a small effect size that may still be biologically relevant. Lastly, although we studied the environmental risk factors prospectively our findings do not indicate a causal relationship with tics; independent replication is necessary using causal designs such as Mendelian randomization (56).

In conclusion, our study made a first step demonstrating that the combination of PRS based on TS cases and pregnancy-related environmental risk factors explained more variance of tics in a general population cohort compared to studying these factors in isolation, pointing to additive effects; thus, suggesting an independent contribution of genes and environment to the development of tics. Our study also suggests mediation effects between environmental variables providing potential clues to underlying pathways. In particular, maternal anxiety/depression and maternal smoking may be associated with a higher number of pregnancy adversities in explaining tics. Continued research efforts in adequately sized prospective clinical and population samples accounting for unmeasured genetic and environmental confounding are needed to uncover how environmental risk factors relate to genetic factors, furthering our understanding of tic disorders.

## Data Availability

The study website contains details of available data through a fully searchable data dictionary: http://www.bristol.ac.uk/alspac/researchers/our-data/.

## ACKNOWLEDGEMENTS

We wish to thank the TS/OCD Working Group of the PGC for providing the summary statistics of the second GWAS of Tourette syndrome. We are also extremely grateful to all the families who took part in this study, the midwives for their help in recruiting them, and the whole ALSPAC team, which includes interviewers, computer and laboratory technicians, clerical workers, research scientists, volunteers, managers, receptionists and nurses.

## CONTRIBUTORS

MA, GAH, PJH, and AD were involved in the organization, design, execution of the research project. MA wrote the first draft of the manuscript, which was critically reviewed by JAT, GAH, PJH, and AD who were also involved in the conception of the research project. All authors have approved the final article.

