## Supplementary material for "Polygenic and environmental determinants of tics in the Avon Longitudinal Study of Parents and Children cohort"

- **Table S1:** Description of the maternal anxiety score
- **Table S2:** Description of the maternal depression score
- **Table S3:** Description of the ALSPAC variables used to construct the cumulative score of adverse pregnancy risk factors
- **Table S4:** Association between the PRS and the environmental predictors in the ALSPAC study
- **Table S5:** Logistic regression models with the predictors maternal anxiety, maternal depression, and the PRS predicting tic presence in the Avon Longitudinal Study of Parents and Children study
- Figure S1: Logistic regression models predicting tic presence in the Avon Longitudinal Study of Parents and Children study (1,2). Area under the curve (AUC) is shown for the models that only contained the PRS as predictor (shown in green) and the models containing the PRS and the environmental variable of interest (shown in red). All models were controlled for sex, maternal socioeconomic status, parity, and maternal smoking.

**Table S1:** Description of the maternal anxiety score

| Item^a^ | Scored as 0 | Scored as 1 | Scored as 2 |
| --- | --- | --- | --- |
| Do you feel upset for no obvious reason? | Not very often / never |  | Very often / often |
| Do you feel strung-up inside? | Not very often / never |  | Very often / often |
| Do you ever have the feeling you are going to pieces? | Not very often / never |  | Very often / often |
| Have you felt as though you might faint? | Never | Not very often | Very often / often |
| Do you feel uneasy and restless? | Never | Not very often | Very often / often |
| Do you worry a lot? | Never | Not very often | Very often / often |
| Do you have bad dreams, which upset you when you wake up? | Never | Not very often | Very often / often |
| Do you sometimes feel panicky | Never |  | Very often / often / not very often |

^a^Maternal anxiety was measured using a self-report of the anxiety subscale of the Crown-Crisp Experiential Index (CCEI; (1–3)) consisting of eight items (range 0-16). Anxiety score was calculated as a sum of all eight items.

**Table S2:** Description of the maternal depression score

| Item^a^ | Scored as 0 | Scored as 1 | Scored as 2 |
| --- | --- | --- | --- |
| Do you feel that life is too much effort? | Never | Not very often | Very often/ often |
| Do you experience long periods of sadness? | Never | Not very often | Very often/ often |
| Do you find yourself needing to cry? | Never | Not very often | Very often/ often |
| Do you have to make a special effort to face up to a crisis or difficulty? | Never | Not very often | Very often/ often |
| Do you regret much of your past behaviour? | Not very often / never |  | Very often /often |
| Do you wake unusually early in the morning? | Not very often / never |  | Very often /often |
| Do you lose the ability to feel sympathy for others? | Not very often / never |  | Very often /often |
| Can you think quickly? | Very often / often |  | Not very often / never |

^a^Maternal depression was measured using a self-report of the anxiety subscale of the Crown-Crisp Experiential Index (CCEI; (1–3)) consisting of eight items (range 0-16). Depression score was calculated as a sum of all eight items.

**Table S3:** Description of the ALSPAC variables used to construct the cumulative score of adverse pregnancy risk factors

| Variable name | Description | Absence of risk (scored as 0) | Presence of risk  (scored as 1) |
| --- | --- | --- | --- |
| Advanced maternal age | Age of mother at birth | <= 40 | >40 |
| Pre-existing medical problem | Complications of a pre-existing medical problem | "No" | "Yes |
| Maternal medication | Medication use in past three months (18 weeks prenatal) | 0 | >= 1 |
|  | Medication use in past three months (32 weeks prenatal) | 0 | >= 1 |
| Maternal diarrhea | Diarrhea during this pregnancy (18 weeks prenatal) | "Not at all" | "Yes in 1-3 months", "Yes four months to now", "Yes both time periods" |
|  | Diarrhea in last three months (32 weeks prenatal) | "No" | "Yes |
|  | Diarrhea in pregnancy from 7 months on | "No" | "Yes |
| Maternal jaundice | Jaundice during this pregnancy (18 weeks prenatal) | "Not at all" | "Yes in 1-3 months", "Yes four months to now", "Yes both time periods" |
|  | Jaundice in last three months (32 weeks prenatal) | "No" | "Yes" |
|  | Jaundice in pregnancy from seven months on | "No" | "Yes" |
| Maternal infections | Any infection in first three months (18 weeks prenatal) | "No" | "Yes |
|  | Any infection (32 weeks prenatal) | "No" | "Yes |
| Morning sickness (excessive vomiting) | Excessive vomiting noted during pregnancy | "No" | "Yes |
| Gestational diabetes/elevated blood sugar | Diabetes in pregnancy | "No glycosuria or diabetes" | "Existing diabetes", "Gestational diabetes", and "Glycosuria" |
| Pre-eclampsia | Pre-eclampsia | "No" | "Yes |

ALSPAC, Avon Longitudinal Study of Parents and Children (REF); CS, cesarean section.

**Table S4:** Association between the PRS and the environmental predictors in the ALSPAC study

| Variable | β | Z | *P* |
| --- | --- | --- | --- |
| Maternal anxiety^a^ | 62.92 | -0.019 | 0.98 |
| Maternal depression^a^ | 51.66 | 1.46 | 0.14 |
| Maternal alcohol | 4.76 | 0.22 | 0.82 |
| Maternal cannabis | -19.41 | -0.21 | 0.84 |
| Maternal smoking | -1.43 | -0.045 | 0.96 |
| Cumulative pregnancy risk score^b^ | 11.54 | 1.02 | 0.31 |

PRS, polygenic risk score; ALSPAC, Avon Longitudinal Study of Parents and Children

^a^Maternal anxiety and chronic maternal depression were measured using the self-rated Crown-Crisp index (3).

^b^A cumulative score of adverse pregnancy risk factors (Table S3).

Table S5: Logistic regression models with the predictors maternal anxiety, maternal depression, and the PRS predicting tic presence in the Avon Longitudinal Study of Parents and Children study

| Model | Without adjustment for maternal smoking | | | | With adjustment for maternal smoking | | | |
| --- | --- | --- | --- | --- | --- | --- | --- | --- |
|  | **N** | **Terms in model** | **R^2^** | **P  (comparison to PRS model)**^a^ | **N** | **Terms in model** | **R^2^** | **P**  **(comparison to PRS model)**^b^ |
| PRS | 4218 | PRS | 0.041 |  |  | PRS, maternal smoking | 0.044 |  |
| Maternal anxiety | 4217 |  |  |  | 4093 |  |  |  |
| Full model |  | PRS, maternal anxiety | 0.053 | 1.4 x 10^-7^ |  | PRS, maternal smoking, maternal anxiety | 0.056 | 2.6 x 10^-7^ |
| Maternal depression | 4215 |  |  |  | 4092 |  |  |  |
| Full model |  | PRS, maternal depression | 0.046 | 0.0007 |  | PRS, maternal smoking, maternal depression | 0.049 | 0.0008 |

PRS, polygenic risk score

^a^Comparison are made with a model that contains the PRS and the confounders sex, maternal socioeconomic status, and parity.

^b^Comparison are made with a model that contains the PRS and the confounders sex, maternal socioeconomic status, parity, and maternal smoking.

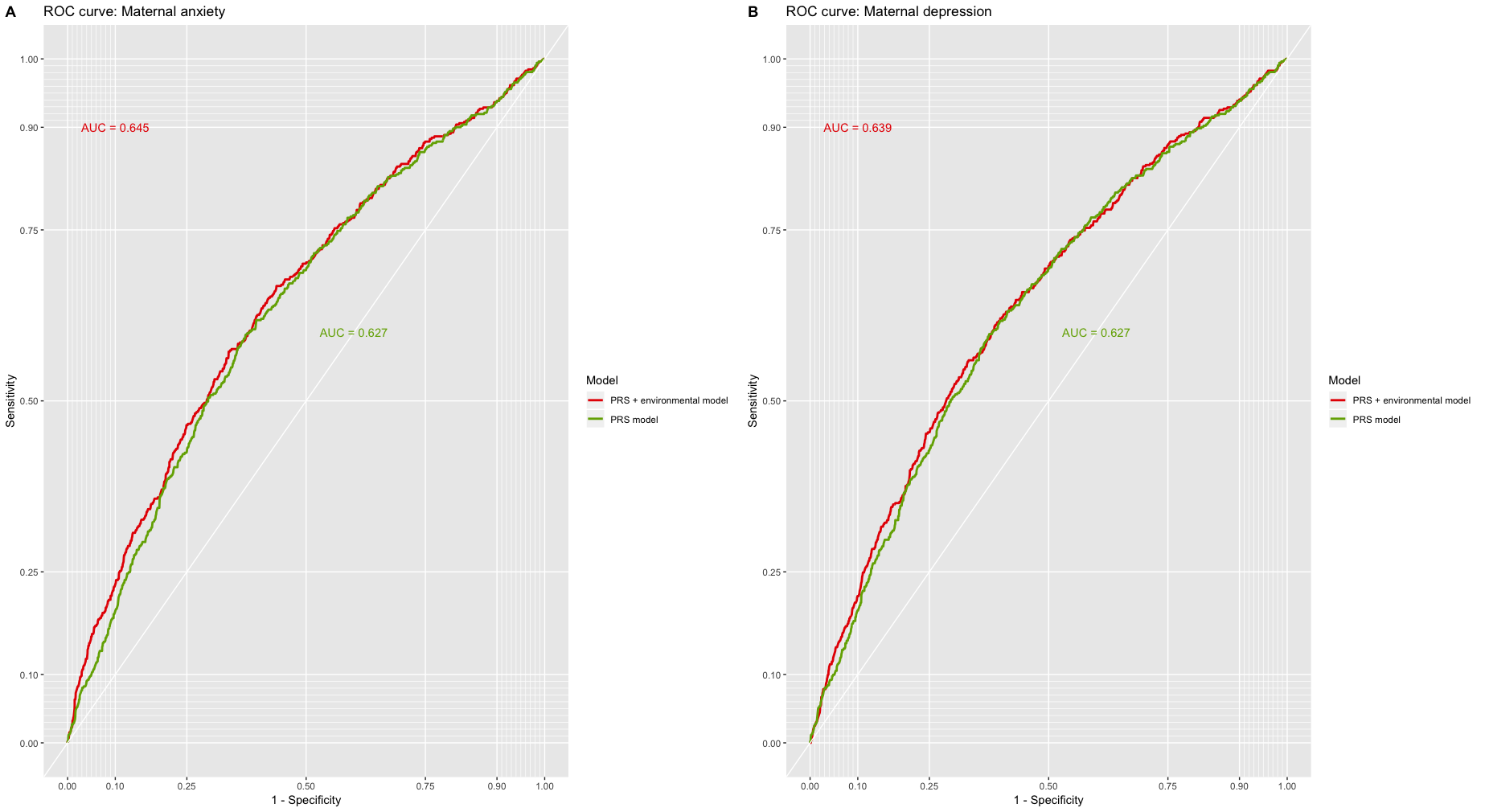

Figure S1: Logistic regression models predicting tic presence in the Avon Longitudinal Study of Parents and Children study (1,2). Area under the curve (AUC) is shown for the models that only contained the PRS as predictor (shown in green) and the models containing the PRS and the environmental variable of interest (shown in red). All models were controlled for sex, maternal socioeconomic status, parity, and maternal smoking.
